# Precise Prediction of COVID-19 in Chest X-Ray Images Using KE Sieve Algorithm

**DOI:** 10.1101/2020.08.13.20174144

**Authors:** S Sai Thejeshwar, Chaitanya Chokkareddy, K Eswaran

## Abstract

The novel coronavirus (COVID-19) pandemic is pressurizing the healthcare systems across the globe and few of them are on the verge of failing. The detection of this virus as early as possible will help in contaminating the spread of it as the virus is mutating itself as fast as possible and currently there are about 4,300 strains of the virus according to the reports. Clinical studies have shown that most of the COVID-19 patients suffer from a lung infection similar to influenza. So, it is possible to diagnose lung infection using imaging techniques. Although a chest computed tomography (CT) scan has been shown to be an effective imaging technique for lung-related disease diagnosis, chest X-ray is more widely available across the hospitals due to its considerably lower cost and faster imaging time than CT scan. The advancements in the area of machine learning and pattern recognition has resulted in intelligent systems that analyze CT Scans or X-ray images and classify between pneumonia and normal patients. This paper proposes KE Sieve Neural Network architecture, which helps in the rapid diagnosis of COVID-19 using chest X-ray images. This architecture is achieving an accuracy of 98.49%. This noninvasive prediction method can assist the doctors in this pandemic and reduce the stress on health care systems.

## I. INTRODUCTION

The novel coronavirus (COVID-19), first appeared in Wuhan, China in December 2019 and has been rapidly spreading across the countries worldwide [5] [6] and subsequently threatening the health of billions of humans and forcing millions of people to stay at home with the majority of countries declaring lockdowns. It has been declared as a pandemic by WHO [11]. The rapid escalation of COVID-19 virus with 2,397,216 confirmed cases and 162,956 confirmed deaths covering 213 countries, areas or territories, (As on 21 April 2020, 2:00 AM CEST) [12] is presenting a huge challenge to the governments across the world, as new cases are being emerged and many new hotspots are evolving on daily basis.

According to the World Health Organisation (WHO), “COVID-19 is the infectious disease caused by the most recently discovered coronavirus. This new virus and disease were unknown before the outbreak began in Wuhan, China, in December 2019.” [9]. This COVID-19 is also called the Severe Acute Respiratory Syndrome CoronaVirus 2 (SARS-CoV-2) [7] [18]. Coronaviruses (CoV) belongs to a family of viruses that cause colds such as the Severe Acute Respiratory Syndrome (SARS-CoV) and the Middle East Respiratory Syndrome (MERS-CoV). This COVID-19 virus is new to human beings and has not been previously identified in humankind. It is said that this virus outbreak is due to the contamination of the virus from animals to humans [8]. According to studies, [8] [9] the SARS-CoV virus is contaminated from musk cats to humans, and the MERS-CoV virus is contaminated from camels to humans. But, the COVID-19 virus is presumed to be contaminated from bats to humans [9]. The COVID-19 virus spreads primarily through droplets of saliva or through cough and sneezing [9]. This led to the transmission of the virus from person to person and caused the rapid spread of the epidemic and had become a pandemic. In this war against COVID-19, all the countries have united together and are fighting against the pandemic. But, this is never about winning at this war, it is about survival. Survival of mankind on earth.

The common symptoms of the disease include fever, tiredness, dry cough. Other symptoms include, shortness of breath, aches and pains, sore throat and very few people had reported diarrhea, nausea, or a runny nose. [9]

The current diagnostic method for COVID-19 is Real-Time Polymerase Chain Reaction (RT-PCR) which uses viral nucleic acid detection in the blood sample. However, there are a limited number of RT-PCR test kits available in most of the countries. Therefore, it is necessary to implement an automatic detection system as a quick alternative diagnosis option to prevent COVID-19 spreading among people. [13] [14] [15]

India, the second-most populous country in the world with 1.2 billion people as of March 2011 [17] and currently has 18,601 confirmed cases and 590 deaths (As on 21 April 2020, 2:00 AM CEST). [12] Given the size of the population these numbers may look small. But, it is a big challenge to the government for the containment of the spread of the disease. According to the Indian Council of Medical Research (ICMR) [18] “A total of 4,01,586 samples from 3,83,985 individuals have been tested as on 19 April 2020, 9 PM 1ST. 17,615 individuals have been confirmed positive among suspected cases and contacts of known positive cases in India.” Are the number of positive cases actually low or they are low because of low testing rate still remains a question. There are many more such countries with low testing rates [19]. But it is essential to increase the testing rate to curb the spread of COVID-19. [15]. Also, some of the eye-opening statistics of India are that 67 percent of the Indian population resides in rural areas and 90 percent of medical imaging facilities are in cities, That makes only 10 percent of imaging facilities spread across small towns in the country [28].

According to the World Health Organization, COVID-19 also opens holes in the lungs like SARS, giving them a “honeycomb-like appearance” [10] [16]. So, in this study, we propose an AI-based pattern recognition system using the KE Sieve Neural Network model [1] [2] for the detection of coronavirus infected patients, pneumonia and healthy patients using chest X-ray radiographs. Also, there are only 3 radiologists per 1,000,000 people in India [28]. So, an automatic screening system would not only help India but also countries across the globe to do a rapid screening and further prevent the spread of the virus. The novelty of this paper is summarized as follows:

1. The proposed model has an end-to-end system without manual human intervention [31].
2. Chest X-ray images are the best tool for the detection of COVID-19.
3. The proposed model classifies between COVID-19, Healthy, and Pneumonia patients.
4. With an increase in the number of training data in the future on a day to day basis, the model can learn as you go.

## II. RELATED WORK

There are few studies on the emergence of COVID-19 virus disease. Fang et al. [20] analyzed computed tomography (CT) scan images using deep neural networks and reported a sensitivity of 98% using 51 COVID-19 patients. Also, Shi et al. [21] collected a large scale COVID-19 CT dataset and developed a machine learning model for COVID-19 screening. Ophir et al. [22] employed deep learning models to detect COVID-19 on CT images. Also, Prabira et al. [23] proposed the detection of COVID-19 using X-ray images based on deep features and SVM. They have extracted the deep features of CNN models and fed them to the SVM classifier. They have obtained 95.38% of accuracy for ResNet50 and Support Vector Machine (SVM). Fei et al. [24], tried to predict COVID-19 patients using “VB-Net” neural network to segment COVID-19 infection regions in CT scans. They obtained dice similarity coefficients of 91.6%±10.0%. Xu et al. [25], proposed a classification model that classifies COVID-19 from viral pneumonia and healthy cases using pulmonary CT images using deep learning techniques.

Their CNN model has yielded the then highest overall accuracy of 86.7 % on CT images. Shuai et al. [26], used CT images to predict COVID-19 cases. They used the inception transfer-learning model to establish the algorithm. They obtained an accuracy of 89.5% with a specificity of 88.0% and a sensitivity of 87.0%.

In some analysis, the normal and pneumonia images have been combined and a binary classification model between COVID-19 and combined classes has been proposed, which is not appropriate as the model will then try to ignore the between-group variance amongst those two classes and the accuracy thus obtained will not be a true measure. It is a better approach to have a tri-class classification model.

The drawback of CT imaging is that typically it takes considerably more time than X-ray imaging. Besides CT scanners may not be available in many regions, making timely COVID-19 screening impossible. In contrast, X-rays are the most common and widely available diagnostic imaging technique [27] and portable in nature. Which makes it a better choice than CT imaging-based detection of COVID-19.

## III. PROPOSED MODEL

The KE Sieve Neural Network [1] [1b] [2] is applied to the Datasets [3] [4] while using features extracted from the transfer learning of pre-trained model weights on the VGG-19 [30] CNN model.

### A. Architecture

The KE Sieve Algorithm [1] is a non-iterative and adopts a new approach, which separates N data points of n-dimension by at least one hyperplane. The number of hyperplanes, *q* needed approximately to separate N data points is the order of *q = log_2_(N)* [3] provided N < 2^n^ and the computational complexity of this algorithm in Big - O asymptotic notation is approximately,

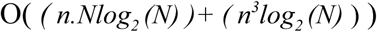

where N is the data points and n is the dimension of space.

1. Consider a set of N train points in an n-dimensional space. Assume this as ‘G’ space. Consider another n-dimensional space as ‘S’, with no points in it. Draw some initial planes in ‘S’ space, where the plane equation is represented as

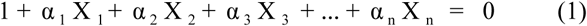
2. To draw the initial planes, collect the random data point pairs of size dimension (n) and substitute them in the above equation (1).
3. Now, calculate the orientation vector (OV) for each point with respect to all the planes in ‘S’ space and then transfer one point at a time from ‘G’ space to ‘S’ space. This Orientation vector gives information about whether the point lies on the positive or negative side of the plane.
4. While transferring from ‘G’ space, OV is calculated and placed in ‘S’ space for the first point. Then from the second point onwards, OV is compared with OV of existing points in ‘S’ space and if OV of the two points differs then the point is placed. Which means the point is separated by plane. If OV of two points matches then point is said to be in the same ‘quadrant’ [1] and the point is not placed. These pairs of points are called neighbors and are collected separately. This pair collection is repeated until it reaches the size of dimension (n). Please refer to Figure 2 below for a 2D representation of ‘S’ space.
5. Now, a new plane is drawn in ‘S’ space by passing through the midpoints of each and every neighbor pair that was collected above. This new single plane is sufficient to separate all the neighbors in ‘S’ space.
6. Also, whenever a new plane is added, orientation vectors (OV) of all the existing points in ‘S’ space are also updated with respect to new hyperplane generated and this process is continued until each and every training point is covered. This ends the process of training the classifier.
7. Now, during predicting the label of a new test point, we compute the dot product of the test point orientation vector with all the train points orientation vectors. Take x% of the maximum value of the dot product results and calculate the Euclidean distance for those train points with the given test point and assign the label of the training point with minimum distance to the test point.

**Figure 1:**
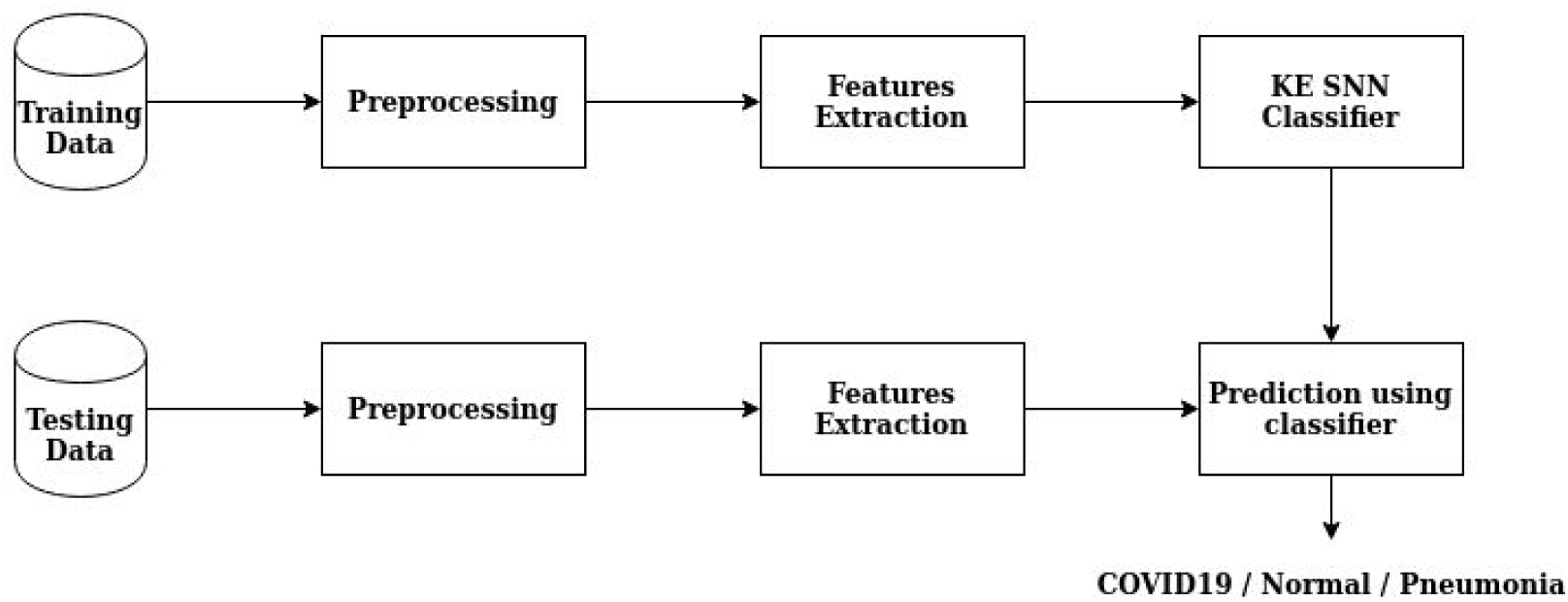
Process Flow Diagram.

**Figure 2:**
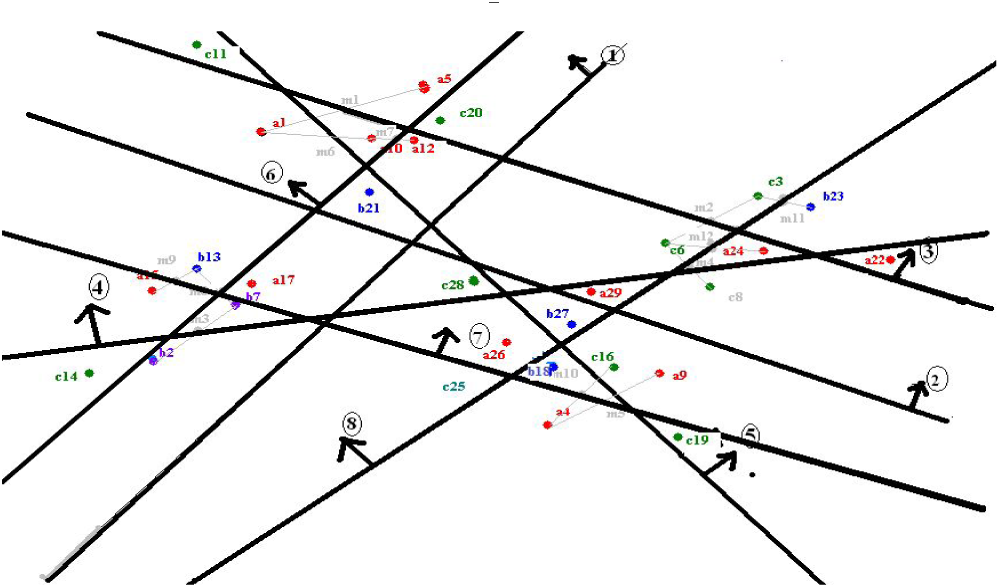
Representation of ‘S’ space point separation in 2D.

This is the only algorithm [1] which has the advantage of incremental learning. That is, if the new points are given for training, the algorithm starts separating these points from where it has previously stopped and OV gets updated accordingly. There is no need to retrain the whole model.

### B. Dataset

The model uses two datasets [3] [4], which has been combined to classify 3 classes as a whole. A total of 108 chest X-ray images of COVID-19 have been taken from this dataset [3]. The other two types of chest X-ray images pneumonia and normal are taken from the Kaggle dataset [4]. Table I shows the distribution of the dataset across the classes. Though the number of COVID-19 images available is too small, it had no effect on the model as a whole as transfer learning-based feature extraction is implemented and SNN [1] mathematically could separate each data point. Figure 3 shows the sample image of the dataset.

**Figure 3:**
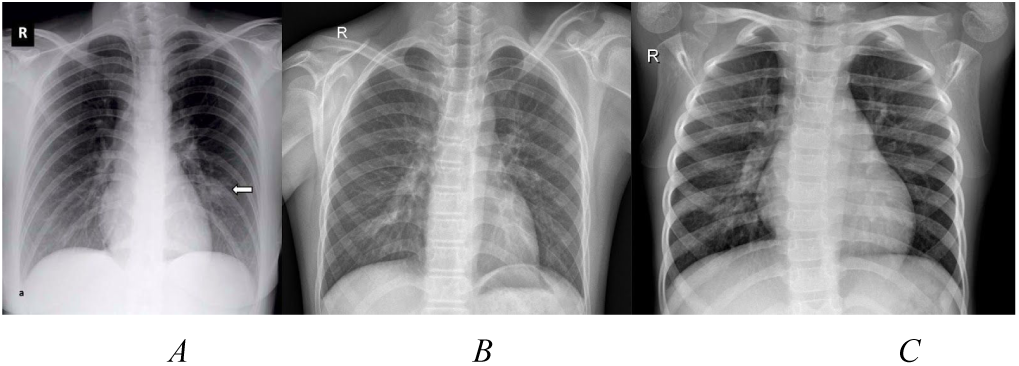
Sample chest X-ray images from the datasets. A - COVID-19; B - Normal; C - Pneumonia.

**Table 1:** Distribution of Dataset.

| Description | COVID-19 | Normal | Pneumonia |
| --- | --- | --- | --- |
| Train Set | 87 | 1,270 | 3,414 |
| Test Set | 21 | 313 | 859 |

### C. Method

We have used the 4,771 train images to train the KE Sieve algorithm (as explained in the *III-A Architecture*) and tested on the 1,193 test images. But, as the classifier expects the inputs to be a real-valued vector, we have done a few preprocessing techniques like converting the image into the standard size of 224 × 224 and then applied the histogram equalisation. Figure 4 below shows the sample output of the preprocessed x-ray images. This preprocessed vector has been fed into the feature extractor of the pre-trained VGG-19 [30] based CNN model. These feature vectors are then fed to the KE Sieve algorithm to consume and separate the train points distinctly. Principal Component Analysis is applied to the feature vector and is plotted in 2 dimensions. Figure 5 below showcases the 2D plot of the feature vector. When this model is tested on the test points it resulted in a new state of the art accuracy of 98.49%. Fig 1 represents the methodology used.

**Figure 4:**
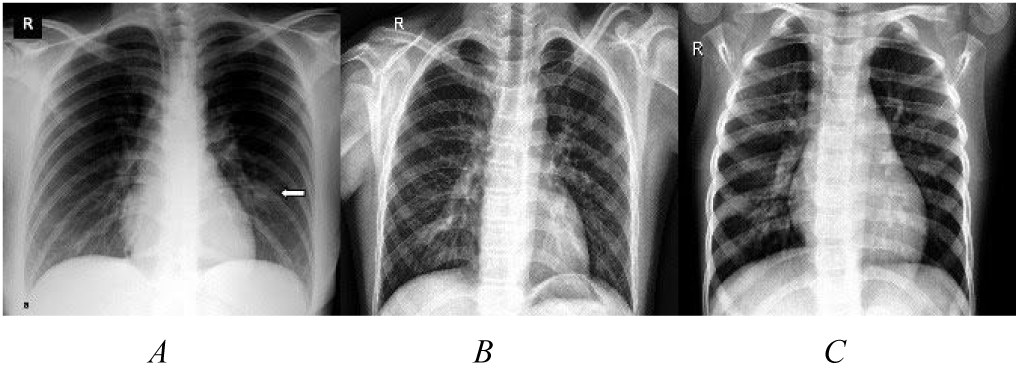
Preprocessed sample chest X-ray images from the datasets. A - COVID-19; B - Normal; C - Pneumonia.

## IV. EXPERIMENTS

In this paper, the input is the image, and the output is the predicted class label. The classification algorithm [1] [2] is implemented in the Python 3 programming language. The environment is the local system with a processor of Intel® Core™ i5-8250U CPU @ 1.60GHz x 8 and a RAM of 8 GB running on Ubuntu OS with no use of GPU.

### A. Evaluation Metrics

In this paper, the primary metric is accuracy for model evaluation. Other metrics like the sensitivity and specificity are also evaluated for the model. The metrics are measured using the confusion matrix (Fig 2). Metrics are defined as,

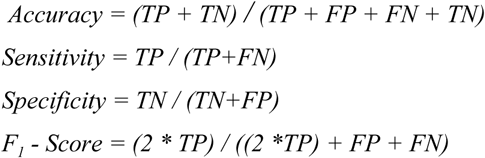

Where, T - True, F - False, P - Positive, N - Negative.

### B. Results

The whole model was rebuilt 10 times, as this helps to keep a check on random initializations.

**Fig 3:**
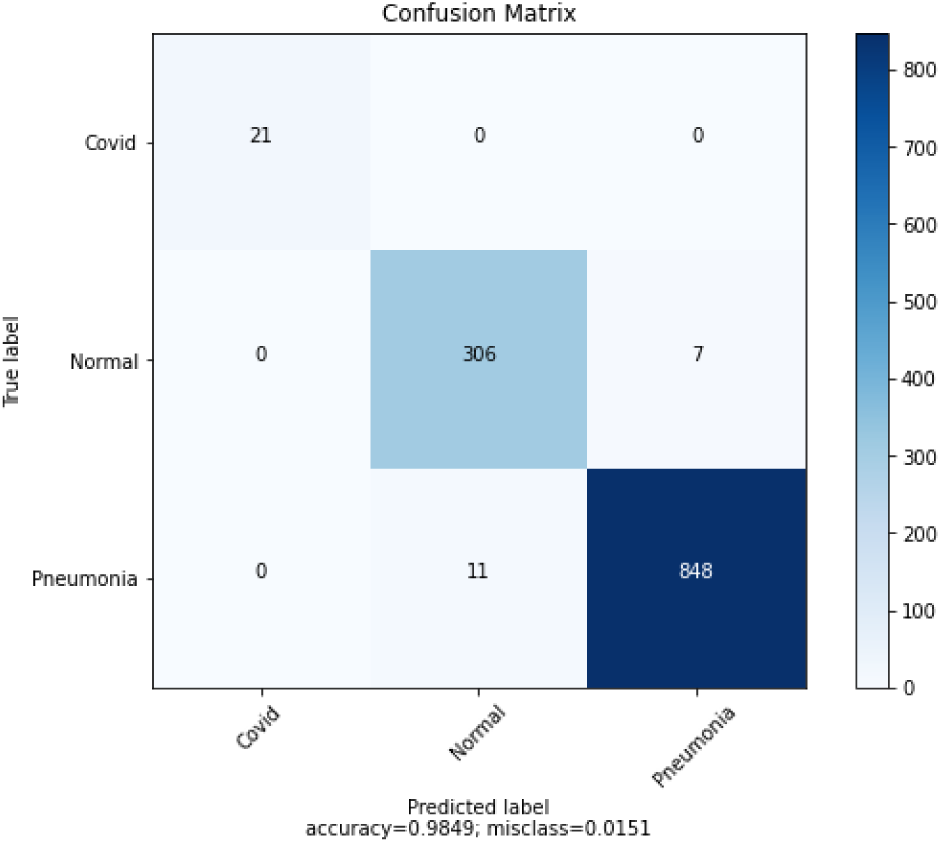
Confusion Matrix.

With the KE Sieve classification approach [1] we just need 18 planes to separate all the train data points in just ~2 seconds. This resulted in an accuracy of 98.49% and sensitivity of 100% and specificity of 100% which is greater than other current models. The other metrics of the model are shown in Table II. From the confusion matrix it is observed that there has been misclassification between Normal and Pneumonia classes. We hope the balanced dataset may overcome this scenario.

**Table -II:**
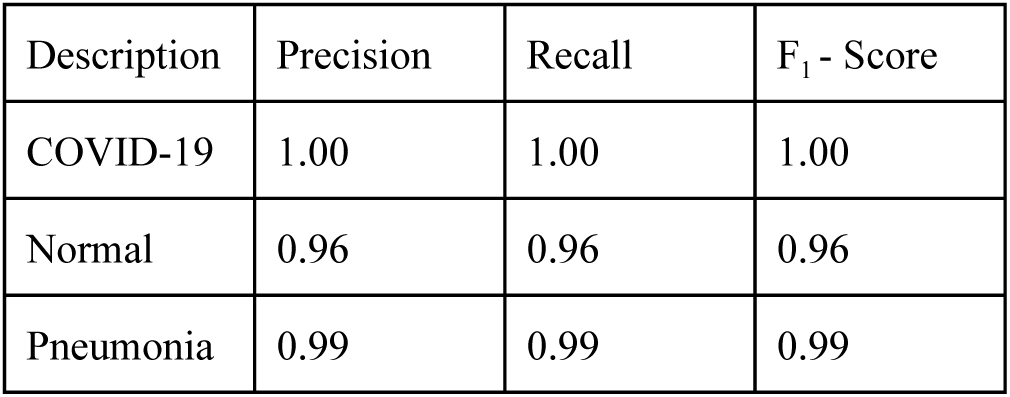
Metrics Report.

The model is built on limited data of COVID-19 is because the disease is new and datasets are not yet available. But we are convinced that with more data also, these results will hold and surely a large number of images would contribute to improving the model, because of the nature of the SNN algorithm [1]. From our experience with KE SNN [1], the more the data points the better the model is as more planes are drawn to separate the data points.

**Figure 5:**
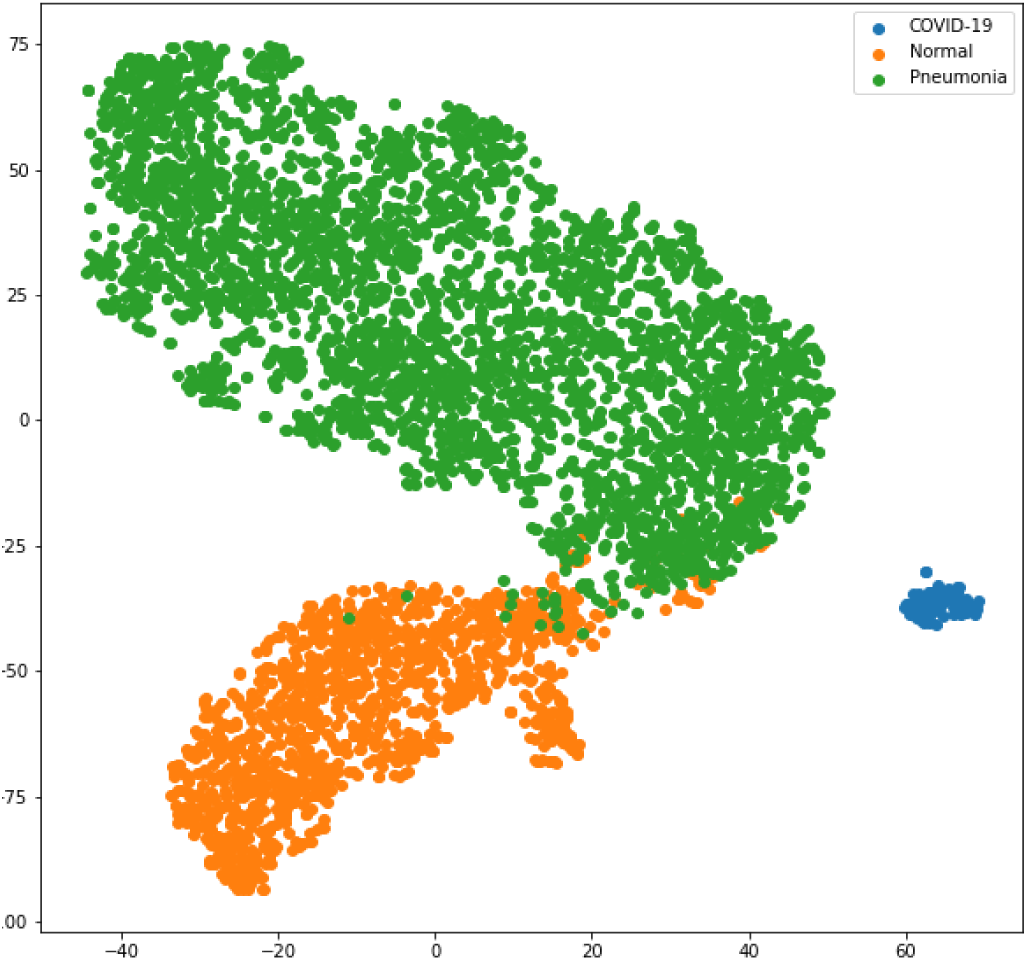
2D Representation of Feature Vectors.

### C. Comparative Study

Our proposed model shows that the classification performance on the dataset [3] [4] is improved and this resulted in higher accuracy when compared with the results of others mentioned in Section II - Related Work.

## V. CONCLUSION

The contribution of this paper is showcasing the effectiveness of this end to end classification model and chest X-ray images are the best tool for the detection of COVID-19 and the proposed model classifies between COVID-19, Healthy, and Pneumonia patients. This model has been deployed as an application [31] and anyone can test it by uploading the X-ray image of the chest.

In our future work, we will explore better ways in feature extraction and classification and also try for the balanced dataset, to ensure the model is not over-fitted or tuned to a specific dataset.

## Data Availability

The dataset used in this research work is available publicly on Github and Kaggle

https://github.com/ieee8023/covid-chestxray-dataset

https://www.kaggle.com/paultimothymooney/chest-xray-pneumonia

